# Comparison of Peripheral Blood Oxygen Saturation Measurements Using Pulse Oximetry at Three Anatomical Sites on Sleep Patients

**DOI:** 10.1101/2022.09.08.22279707

**Authors:** Kellie R. Strickland, Chris J. Brown, Leigh Wilks, Peter K. Dunn, Mark A. Holmes

## Abstract

**Background and Objective:** Accurate measurements of peripheral blood oxygen saturation (SpO_2_) are important in evaluating sleep patients with frequent desaturations due to pathological breathing events. This project compared synchronised SpO_2_ measurements at the finger, forehead and toe of patients undergoing Type 1 polysomnography (PSG) to evaluate potential SpO_2_ variability across the sites.

**Methods:** Pulse oximetry SpO_2_ measurements were simultaneously and continuously recorded for 41 sleep patients at the finger, forehead and toe, and synchronised with PSG data. Recordings were scored for desaturations of ≤ 3% (peak to trough, lasting ≤10 seconds), signal dropouts, and artefact occurrences. Forehead and toe SpO_2_ measurements were compared against the finger as the standard PSG oximetry site.

**Results:** Differences between anatomical sites for mean SpO_2_, mean number of SpO_2_ desaturations per hour, and time spent below an SpO_2_ level of 95% during total sleep time were significant (*P* < 0.01). The forehead pulse oximeter had the highest mean SpO_2_, least number of SpO_2_ desaturations per hour, and experienced the least number of artefact occurrences. Dropouts were lowest for the forehead and toe pulse oximeters.

**Conclusion:** Differences between SpO_2_ measurements, dropouts and artefact occurrences at the finger, forehead and toe may have diagnostic and prognostic implications for sleep patients. The differences in SpO2 measurements may be attributed to variability in perfusion of the extremities and core during sudden oscillating blood pressure changes associated with breathing events. Further research is required to determine which anatomical site correlates closest to arterial oxygenation for pulse oximetry in sleep patients.

**BRIEF SUMMARY:** *Study rationale:* An absence of research investigating anatomical site location for pulse oximetry during overnight polysomnography exists. Our study was performed to fill this gap, as accurate pulse oximetry measurements are key for the diagnosis, treatment and monitoring of sleep patients; a patient cohort where SpO_2_ desaturations are recurrent.

*Study impact:* Our findings demonstrate there are significant differences between finger, forehead and toe pulse oximetry measurements, particularly SpO_2_ desaturations per hour, which may have diagnostic and clinical implications. This research is applicable and important to not only sleep physicians and scientists, but also other disciplines where continuous SpO_2_ monitoring is required.

## INTRODUCTION

Pulse oximetry is a non-invasive technique used in sleep diagnostics to assess patient blood oxygenation (SpO_2_) levels and for detecting hypoxaemia ^1-4^. A sleep study, also known as polysomnography (PSG), is used to diagnose obstructive sleep apnoea (OSA) and other sleep disorders. Patients undergoing PSG for the investigation of sleep disordered breathing often demonstrate significant and numerous blood oxygen desaturations that are a hallmark characteristic of OSA ^2, 3, 5^. PSG provides a comprehensive multi-parametric recording of the physiological changes that occur during sleep and detects pathological breathing events ^2,3^. Alternatively, an overnight oximetry study either at home or in hospital can be conducted as a surrogate investigation or initial screening process to assess sleep disorders. Both types of sleep studies require continuous and accurate monitoring of patient SpO_2_ levels ^2,3^.

Pulse oximetry is limited by factors such as peripheral perfusion, motion artefact and vasoconstriction ^1, 6-10^, which may be more prevalent when SpO_2_ is measured at different anatomical locations. Previous studies have reported that forehead pulse oximeters provided a more accurate and consistent SpO_2_ reading than finger pulse oximeters in patients undergoing post-anaesthesia care, ambulance transportation and respiratory assessment ^11-15^. Anatomical characteristics of the forehead, such as a higher level of tissue perfusion, fewer occurrences of vasoconstriction, and less risk of motion artefacts, may have contributed to the observed difference in SpO_2_ measurements ^11-15^. Two studies reported that SpO_2_ measurements with earlobe pulse oximeters in patients with chronic obstructive pulmonary disease (COPD) overestimated arterial blood oxygen saturation (SaO_2_) to such an extent that the earlobe was deemed inappropriate for evaluating oxygen administration and overall clinical decision-making ^16,17^.

Limited research exists comparing simultaneous SpO_2_ measurements on patients undergoing PSG using pulse oximeters at different anatomical sites. The failure of a pulse oximeter to detect an oxygen desaturation during a PSG means that the corresponding breathing event may not be scored, and the overall severity of the patient’s clinical condition may be underestimated or treatment under-titrated ^2,3^. The aim of this cross-sectional study was to compare simultaneous, synchronised SpO_2_ measurements obtained with pulse oximeters located at three anatomical sites (finger, forehead and toe) on patients undergoing Type 1 PSG. The mean SpO_2_, number and magnitude of SpO_2_ desaturations per hour, number of signal dropouts, and occurrence of artefactual readings were compared for pulse oximetry measurements at the three anatomical sites.

## METHODS

Ethical approval was granted by The Prince Charles Hospital Human Research Ethics Committee (HREC/18/QPCH/179), with subsequent authorisation to conduct the project at the Sleep Assessment Unit at the Sunshine Coast University Hospital (SCUH) (QSC/40666). Patients undergoing Type 1 PSG were recruited from August 2018 to December 2018 by non-probability convenience sampling. Exclusion criteria included: any form of skin infection at the three anatomical sites; impaired nail quality; any medical condition known to affect pulse oximeter accuracy; and an inability to participate in Type 1 PSG. Patients recruited for the project completed a self-reported, medical history checklist consisting of 38 common medical conditions.

Patients had a pulse oximeter attached at their finger, forehead and toe, with the photo-emitter and detector placed as per each manufacturer’s specifications. A NONIN pulse oximeter with soft-tip finger probe was placed on the middle finger as per standard practice for Type 1 PSG. A Masimo pulse oximeter integrated into a Sensor 92 on a Radiometer TCM4 TOSCA monitoring system was used on the forehead and set to 43°C. A Masimo Radical 7 pulse oximeter with disposable probe was placed on the best fitting toe. Oximetry outputs were recorded at a sample rate of 16Hz. The three oximeters were recorded simultaneously in conjunction with other routine electrode channels and synchronized with other PSG parameters. All electrodes were monitored continuously overnight by a sleep scientist in the Sleep Assessment Unit. Pulse oximetry data was scored according to strict rules (Fig. 1) to differentiate the commencement and termination of SpO_2_ desaturation events. To avoid scorer bias, pulse oximeter data was scored without the anatomical site label. Each patient’s full PSG was scored according to the American Academy of Sleep Medicine (AASM) manual in conjunction with Australasian Sleep Technologists Association (ASTA) and Australasian Sleep Association (ASA) commentary ^2,3^.

**Figure 1:**
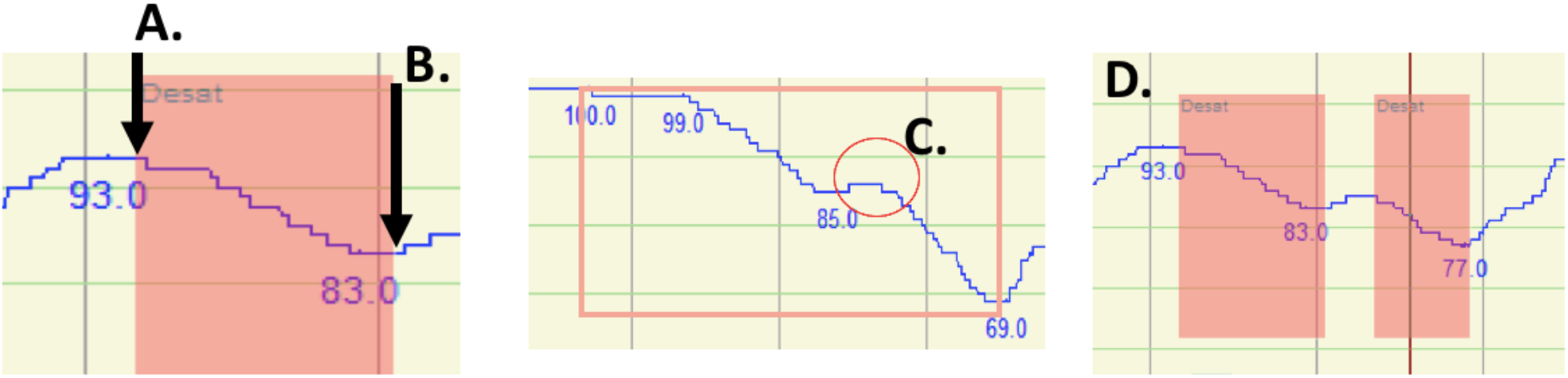
SpO_2_ desaturation events were measured precisely from peak (A.) to end of trough (B.). When an increase in SpO_2_ of less than 3% was followed by a continued SpO_2_ decrease during a desaturation (C.) that did not correlate with an increase in patient respiration, the increase was disregarded and one desaturation event was scored. When the increase was 3% or greater, then two individual desaturations were scored (D.). If the SpO_2_ trace dropped out completely or had substantial artefact occurrences during a suspected desaturation, the event was not scored as a desaturation but as a dropout or artefact occurrence because an accurate peak and trough was not identifiable.

De-identified patient data including PSG results, pulse oximetry measurements, age, gender, height, weight and body mass index (BMI) were transferred to SPSS® (Version 24, IBM) for statistical analysis. Descriptive statistics were used to quality check and summarise the data, and reported as mean (SD). Parametric tests were used for inference due to the sample size (n=41). The assumptions for all statistical tests performed were met. Levene’s test for equality of variances was undertaken before conducting all unpaired t-tests. A Repeated Measures Analysis of Variance (RMANOVA) was used to determine significant differences between the three pulse oximeters for all SpO_2_ measurements. The relationship between oximeter values were determined using Pearson’s correlation coefficient (r). Bland-Altman plots were used to complement the correlation analysis, and to show the level of agreement between the oximeters at the different anatomical sites, using the finger as the standard PSG oximetry site ^18, 19^. Statistical significance was chosen at *P* < 0.05.

## RESULTS

Table 1 summarises the demographic attributes of the Type 1 PSG patients (n=41) recruited for the project. The patients underwent one of the following: diagnostic study (39%, n=16), split diagnostic/Continuous Positive Airway Pressure (CPAP) study (19.5%, n=8), CPAP titration study (36.6%, n=15) or bilevel positive airway pressure titration study (4.9%, n=2). Most patients (91%, n=30) had sleep disordered breathing, including a mild OSA (42.4%, n=14) or moderate OSA (24.2%, n=8). Half the patients (51.2%, n=21) had four or more medical conditions, of which 17% had ten or more conditions. Many patients reported a cardiac history (58.5%, n=24) or respiratory condition (40%, n=17). Other reported conditions were anxiety (43.9%, n=18), depression (36.6%, n=15), gastric reflux (41.5%, n=17) and diabetes (29.3%, n=12).

**Table 1.** Demographic attributes of the recruited patients undergoing PSG (n=41).

| Characteristic | Males | Females | Total | 95% CI | Range |
| --- | --- | --- | --- | --- | --- |
|  | (n = 23) | (n = 18) | (n = 41) |  |  |
| Age (years) | 61.1 (18.01) | 64.2 (9.82) | 62.1 (14.57) | 57.1 to 67.0 | 18 – 84 |
| Height (cm)* | 173.3 (5.24) | 161.8 (6.05) | 168.1 (8.40) | 165.4 to 170.8 | 150 – 185 |
| Weight (kg) | 96.2 (14.57) | 88.4 (20.44) | 94.2 (20.61) | 87.2 to 101.1 | 62 – 162 |
| BMI (kg/m<sup>2</sup>) | 31.8 (4.40) | 33.8 (7.56) | 33.2 (6.90) | 30.9 to 35.6 | 23.3 – 55.4 |
| Neck circumference (cm) | 41.0 (4.26) | 38.3 (3.65) | 40.0 (1.38) | 38.5 to 41.5 | 33 - 50 |
| ESS | 5.2 (3.86) | 8.8 (5.30) | 7.7 (5.40) | 5.7 to 9.6 | 0 – 21 |
| AHI Mean<sup>1</sup> | 19.5 (13.23) | 22.5 (25.28) | 19.9 (19.5) | 12.86 to 26.9 | 1 – 85 |
| Baseline SpO<sub>2</sub> (%)<sup>2</sup> | 96.6 (1.55) | 95.8 (1.75) | 96.2 (1.60) | 95.6 to 96.8 | 92 – 99 |

Data are reported as either mean (SD), 95% confidence interval (CI) or as absolute values.
Statistical significance between males and females was measured using an unpaired t-test;
\*significant level at $P < 0.05$
<sup>1</sup>Eight diagnostic/CPAP split studies were not included in the AHI calculation due to actively changing AHI from direct therapeutic intervention during an overnight study.
<sup>2</sup>Measured by finger pulse oximetry as the standard PSG site.
Note: PSG: Polysomnography; BMI: Body Mass Index; ESS: Epworth Sleepiness Scale; AHI: Apnoea Hypopnea Index; CPAP: Continuous Positive Airway Pressure.

Mean SpO_2_ during total sleep time at the forehead was significantly higher than toe oximetry and finger oximetry measurements (RMANOVA: F (1.245, 48.551) = 38.782, *P* < 0.01; Table 2). The percentage difference in both mean magnitude of SpO_2_ desaturations and largest SpO_2_ desaturation during total sleep time was smallest between the finger and forehead pulse oximeters. While there was no significant difference between the three pulse oximeters for the largest SpO_2_ desaturation during total sleep time (RMANOVA: F (1.333, 49.311) = 2.972, *P* = 0.08), the mean number of SpO_2_ desaturations detected per hour were significantly different (RMANOVA: F (1.427, 57.066) = 14.891, *P* < 0.01, Table 2). The finger pulse oximeter detected the most SpO_2_ desaturations per hour and the forehead pulse oximeter detected the least number. A high degree of variability existed with all pulse oximeters for the detection of the mean number of SpO_2_ desaturations per hour.

**Table 2.** Comparison of frequently reported SpO_2_ indices collected during PSG.

| Variable | Mean (SD) | 95% CI | Range |
| --- | --- | --- | --- |
| **Difference in mean SpO<sub>2</sub> during total sleep time (%)** |  |  |  |
| Finger - Forehead | -1.4 (1.07)<sup>a</sup> | -1.7 to -1.1 | -3 to 4 |
| Finger - Toe | -1.0 (1.08)<sup>b</sup> | -1.4 to -0.7 | -3 to 2 |
| Forehead - Toe | 0.3 (0.97)<sup>c</sup> | -0.0 to 0.6 | -2 to 2 |
| **Difference in mean magnitude of SpO<sub>2</sub> desaturations (%)** |  |  |  |
| Finger - Forehead | -0.01 (0.982)<sup>a</sup> | -0.3 to 0.3 | -4 to 3 |
| Finger - Toe | -0.37 (0.999)<sup>b</sup> | -0.7 to -0.1 | -5 to 1 |
| Forehead - Toe | -0.39 (0.721)<sup>a,b</sup> | -0.6 to -0.1 | -3 to 1 |
| **Difference in largest SpO<sub>2</sub> desaturation (%)** |  |  |  |
| Finger - Forehead | -0.31 (2.48) | -1.1 to 0.5 | -8 to 4 |
| Finger - Toe | -1.42 (2.97) | -2.4 to -0.4 | -13 to 3 |
| Forehead - Toe | -1.18 (2.61) | -1.9 to -0.3 | -13 to 2 |
| **Mean number of SpO<sub>2</sub> desaturations per hour (absolute values)** |  |  |  |
| Finger | 19.2 (23.37)<sup>a</sup> | 11.8 – 26.6 | 0 – 108.24 |
| Forehead | 11.8 (14.21)<sup>b</sup> | 7.3 – 16.2 | 0 – 60.92 |
| Toe | 14.0 (18.29)<sup>c</sup> | 8.3 – 19.8 | 0 – 86.88 |

Data are reported as mean (SD) with a 95% confidence interval (CI).
Statistical significance was measured using a repeated measures ANOVA;
<sup>a,b,c</sup> Values followed by different letters within columns differ significantly ( $P < 0.05$ ). Values with the same letter do not differ significantly.
*Note:* PSG: Polysomnography.

The mean number of SpO_2_ desaturations per hour during total sleep time showed a strongly positive and significant relationship between the finger and forehead pulse oximeters (R^2^ = 0.937, *P* < 0.01, Fig. 2A) and the finger and toe pulse oximeters (R^2^ = 0.942, *P* < 0.01, Fig. 2C). A Bland-Altman plot confirmed that the mean number of SpO_2_ desaturations detected per hour by the finger pulse oximeter was higher than both the forehead and toe oximeters, with wide limits of agreement of -14.56 to 29.42% (Fig. 2B) and -11.89 to 22.20% (Fig. 2D), respectively. The mean number of SpO_2_ desaturations per hour were more likely to fall outside the limits of agreement when values were greater than 20 desaturations per hour.

**Figure 2:**
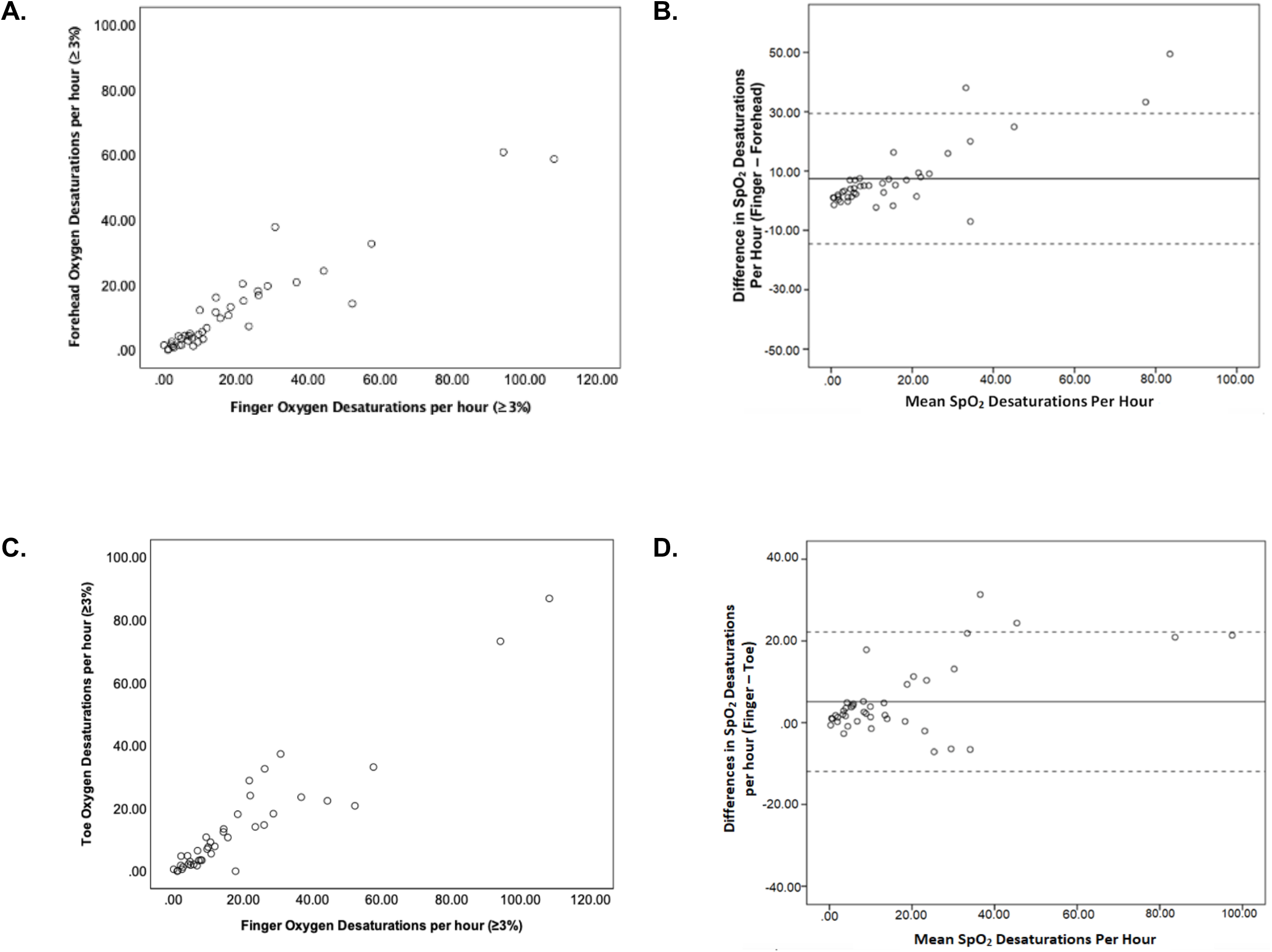
Relationship between finger versus forehead and finger versus toe for number of SpO_2_ desaturations detected per hour during total sleep time (n=41), described by Pearson correlations (A. & C.) and Bland-Altman plots (B. & D.). Pearson’s r was 0.929 (95% CI 0.869 to 09.62; *P <*0.001) for finger versus forehead (A.), and 0.948 (95% CI 0.902 to 0.973; *P* < 0.001) for finger versus toe (B.). For Bland-Altman plots, the solid lines indicate the mean difference in number of SpO_2_ desaturations per hour between the pulse oximeters, and the dashed lines indicates the limits of agreement.

A significant difference was observed between the three pulse oximeters for the time spent at an SpO_2_ below 95% during total sleep time for the 41 patients (RMANOVA: F (1.178, 45.933) = 24.430, *P* < 0.01, Table 3). The finger pulse oximeter spent the greatest time below an SpO_2_ of 95% and the forehead pulse oximeter the least time. For one patient, the finger pulse oximeter spent over 3 hours longer below an SpO_2_ level of 95% than the forehead and toe pulse oximeter. While SpO_2_ dropped below 88% during total sleep time for 11 patients (Table 3), the difference between pulse oximeters was not significant (RMANOVA: F (1.051, 10.506) = 3.144, *P* = 0.104).

**Table 3.** Differences in the total time spent at an SpO_2_ level below 95% and 88% during PSG.

| Variable | Mean (SD) | 95% CI | Range |
| --- | --- | --- | --- |
| **Difference in total time (minutes) spent at an SpO<sub>2</sub> below 95%** |  |  |  |
| **(n = 41)** |  |  |  |
| Finger - Forehead | 60.68 (61.3)<sup>a</sup> | 41.3 to 80.0 | -71 to 198 |
| Finger - Toe | 32.53 (59.6)<sup>b</sup> | 13.8 to 51.2 | -163 to 174 |
| Forehead - Toe | -28.73 (64.9)<sup>c</sup> | -49.5 to -8.0 | -199 to 73 |
| **Difference in total time (minutes) spent at an SpO<sub>2</sub> below 88%** |  |  |  |
| **(n = 11)** |  |  |  |
| Finger - Forehead | 7.82 (11.57) | 0.1 to 15.6 | -5 to 33 |
| Finger - Toe | -1.82 (13.81) | -11.1 to 7.5 | -40 to 16 |
| Forehead - Toe | -2.65 (11.75) | -6.4 to 1.1 | -73 to 4 |

Data are reported as mean (SD) with a 95% confidence interval (CI).
Statistical significance was measured using a repeated measures ANOVA;
<sup>a b c</sup> Values followed by different letters within columns differ significantly ( $P < 0.05$ ).
Note: PSG: Polysomnography.

Table 4 shows the finger pulse oximeter had a significantly higher mean number of signal dropouts per patient (RMANOVA: F (1.086, 41.336) = 12.544, *P* < 0.01) and dropouts per hour (F (1.095, 42.718) = 12.804, P < 0.01) than the forehead and toe pulse oximeters during total report time. Furthermore, the forehead pulse oximeter had significantly lower mean occurrences of artefact (RMANOVA: F (1.218, 47.487) = 4.594, *P* = 0.030) and rate of artefact per hour (RMANOVA: F (1.237, 48.235) = 4.969, *P* = 0.024) than the finger and toe pulse oximeters during total report time.

**Table 4.**
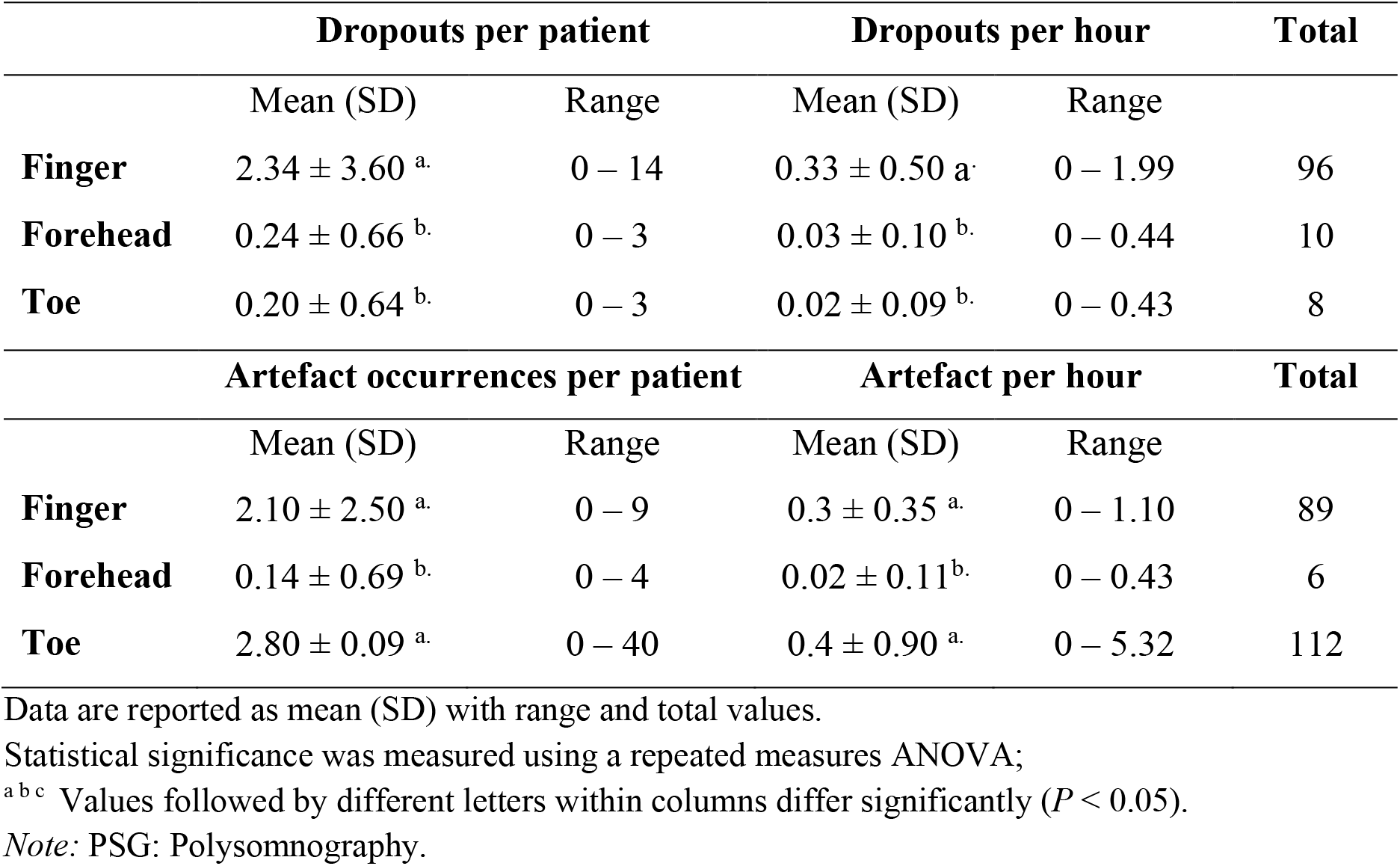
Comparison of signal dropouts and artefact occurrences during PSG.

|  | Dropouts per patient |  | Dropouts per hour |  | Total |
| --- | --- | --- | --- | --- | --- |
|  | Mean (SD) | Range | Mean (SD) | Range |  |
| **Finger** | $2.34 \pm 3.60^a$ | 0 – 14 | $0.33 \pm 0.50^a$ | 0 – 1.99 | 96 |
| **Forehead** | $0.24 \pm 0.66^b$ | 0 – 3 | $0.03 \pm 0.10^b$ | 0 – 0.44 | 10 |
| **Toe** | $0.20 \pm 0.64^b$ | 0 – 3 | $0.02 \pm 0.09^b$ | 0 – 0.43 | 8 |
|  | Artefact occurrences per patient |  | Artefact per hour |  | Total |
|  | Mean (SD) | Range | Mean (SD) | Range |  |
| **Finger** | $2.10 \pm 2.50^a$ | 0 – 9 | $0.3 \pm 0.35^a$ | 0 – 1.10 | 89 |
| **Forehead** | $0.14 \pm 0.69^b$ | 0 – 4 | $0.02 \pm 0.11^b$ | 0 – 0.43 | 6 |
| **Toe** | $2.80 \pm 0.09^a$ | 0 – 40 | $0.4 \pm 0.90^a$ | 0 – 5.32 | 112 |

Data are reported as mean (SD) with range and total values.
Statistical significance was measured using a repeated measures ANOVA;
<sup>a b c</sup> Values followed by different letters within columns differ significantly ( $P < 0.05$ ).
Note: PSG: Polysomnography.

## DISCUSSION

Our study found differences between SpO_2_ values measured with pulse oximeters positioned at the finger, forehead and toe of patients undergoing Type 1 PSG, which is consistent with previous studies of patients in post anaesthesia care, intensive care units, ambulatory transportation, and with chronic obstructive pulmonary disease ^11-17, 20-26^. The finger pulse oximeter, which is the standard PSG oximetry site, recorded the lowest mean SpO_2_ and highest mean number of SpO_2_ desaturations per hour during total sleep time. However, the finger pulse oximeter also experienced the highest number of dropouts per patient and per hour as compared to the forehead and toe pulse oximeters. Artefact occurrences were highest for both the finger and toe pulse oximeters. Such differences between oximeters for the continuous monitoring of SpO_2_ during total sleep time may have diagnostic and prognostic implications for patients undergoing PSG, including the assessment of OSA severity with the AHI and other oxygen saturation indices ^2-4^.

Several studies have reported that forehead oximetry recorded a higher mean SpO_2_ and had better agreement with fewer outliers than finger oximetry when compared to SaO_2_ for respiratory and intensive care patients ^11, 12, 15^. In contrast, Sohila *et al*. found the forehead oximeter recorded a lower mean SpO_2_ than finger, toe and earlobe oximeters ^26^. While our study showed differences for mean SpO_2_ across total sleep time between the three oximetry sites, these differences were within acceptable limits of agreement (± 2%) set by manufacturers for their pulse oximeters ^6, 27^ and are unlikely to be clinically significant.

Patients with OSA experience frequent blood oxygen saturations associated with pathological breathing events ^5^. We found poor agreement between the three oximeter sites when the frequency of SpO_2_ desaturations per hour was >20, with some SpO_2_ values falling outside the wide limits of agreement (Figs. 2A & 2C). This could influence patient diagnosis and classification of OSA severity, and result in suboptimal treatment, such as the prescription of inadequate CPAP pressures ^2, 3^. For instance, the classification of OSA severity for one of the patients in our study was graded as severe with finger oximetry (55 SpO_2_ desaturations per hr), moderate with toe oximetry (21 SpO_2_ desaturations per hr), and mild with forehead oximetry (14 SpO_2_ desaturations per hr). Differences in frequency of SpO_2_ desaturations detected per hour by each oximeter is concerning for home sleep apnoea testing and overnight oximetry screens that depend on oximetry measurements alone and lack informative supporting PSG parameters^35^.

The vigorous cycle of sudden oscilating blood pressure and perfusion changes due to obstructive respiratory events may have different effects on oximetry readings at the core and extremities. Vasoconstriction and vasodilation can cause oximeters to overestimate or underestimate SaO_2_ in healthy volunteers ^19, 10, 28, 29^. Furthermore, the degree of hypoxaemia experienced during obstructive respiratory events may be less severe at the forehead than extremities because of the protective cerebrovascular autoregulation mechanism ^30-32^, thereby explaining why the forehead oximeter detected the least number of SpO_2_ desaturations per hour. The clinical significance of our findings requires further investigation.

The finger oximeter spent the most time below an SpO_2_ of 95% (Table 3), with one patient recording over 3-hours longer below an SpO_2_ of 95% than the forehead and toe oximeters. Since the baseline SpO_2_ was approximately 95% for some patients, small variations in SpO_2_ above and below this clinical limit are likely to have contributed to the poor agreement between oximeters. In contrast, the difference in time spent below 88% rarely exceeded 10 minutes, suggesting good agreement between oximeters at this clinical limit. While this difference is insignificant during a total sleep period, it could be problematic in other clinical settings such as ICU where measurement time of SpO_2_ is critical ^33, 34^.

Since SpO_2_ measurements are crucial for apnoea and hypopnea scoring in patients undergoing PSG, signal quality of pulse oximeters at each anatomical site is imperative ^2, 3, 26,^. In our study, the forehead oximeter had better signal stability and reliability than the finger and toe oximeters, with only ten dropouts and six artefact occurrences being recorded across 280 hours of sleep data for 41 patients (Table 4). Similar findings were reported for forehead oximetry in patients undergoing ambulance transportation and postanaesthesia care ^13, 14^. This suggests that signals from pulse oximeters placed on the extremities are less stable and reliable than those placed more central to the core. Increased risk of motion artefact, vasoconstriction, and decreased perfusion at the extremities, all of which can weaken the photoplethysmographic signal ^7, 9, 10^, may contribute to oximetry instability at the three anatomical sites selected for our study.

## LIMITATIONS

Our research study had several limitations. First, comparing the pulse oximetry SpO_2_ measurements against arterial blood gas (ABG) measurements to ascertain which pulse oximeter recorded values closest to SaO_2_ values was not possible. ABG samples taken from patients undergoing PSG are impractical because they are invasive and painful, and not recommended for standard clinic practice ^2, 3^. Secondly, the finger pulse oximeter was manufactured by a different company to the oximeters attached to patients’ foreheads and toes. However, pulse oximeters utilise similar theoretical principles and algorithms and should not vary substantially to influence SpO_2_ measurements. Finally, our cross-sectional study included a small sample of sleep patients with a range of medical conditions. The study exclusion criteria sought to minimise confounders of SpO_2_ measurements at the three anatomical sites. Nevertheless, SpO_2_ measurements at the anatomical sites may have been affected by individual pathologies in unknown ways, which requires further investigation.

## CONCLUSION

Our study has compared SpO_2_ measurements using pulse oximetry at three anatomical sites (finger, forehead, toe) on patients undergoing Type 1 PSG. The forehead pulse oximeter recorded the highest mean SpO_2_ level, detected the least SpO_2_ desaturations, and had the most stable and reliable signal in relation to dropouts and artefact occurrences. The differences in SpO_2_ measurements from the three oximeter sites may be attributed to variability in perfusion between the extremities and core during sudden oscillating blood pressure changes associated with obstructive breathing events. We recommend that careful consideration be given to the interpretation of SpO_2_ measurements from different oximetry sites for the diagnosis and treatment of patients undergoing PSG. Clearly, further research is required to determine with confidence which pulse oximetry site most accurately reflects changes in SaO_2_ levels for patients undergoing sleep studies.

## Supporting information

STROBE checklist

ICMJE disclosure form

## Data Availability

Patient data supporting the findings of this project are available from the Sunshine Coast Hospital and Health Service (SCHHS). Restrictions apply to the availability of these data, which were used under license for this project. Data are available upon request.

## DATA AVAILABILITY STATEMENT

Patient data supporting the findings of this project are available from the Sunshine Coast Hospital and Health Service (SCHHS). Restrictions apply to the availability of these data, which were used under license for this project. Data are available from Chris Brown with permission of the SCHHS.

## ACKNOWLEDGEMENT

The authors acknowledge the SCUH Sleep Assessment Unit and SCHHS for the use of their clinical facilities and assistance from their sleep scientists. This project received no external funding.

## ABBREVIATIONS

AASM: American Academy of Sleep Medicine
ABG: Arterial Blood Gas
AHI: Apnoea Hypopnea Index
ATSA: Australasian Sleep Technologists Association
ASA: Australasian Sleep Association
BMI: Body Mass Index
COPD: Chronic Obstructive Pulmonary Disease
CPAP: Continuous Positive Airway Pressure
ICU: Intensive Care Unit
OSA: Obstructive Sleep Apnoea PSG: Polysomnography
SaO_2_: Arterial blood oxygen saturation
SpO_2_: Peripheral blood oxygen saturation

## Notes

This research project was performed within the Sleep Assessment Unit at the Sunshine Coast University Hospital. All authors have seen and approved the manuscript and agree with its submission to MedRxiv. This project received no external funding, and all authors have no conflicts of interest to declare.

### Competing Interest Statement

The authors have declared no competing interest.

### Author Declarations

The Prince Charles Hospital Human Research Ethics Committee(EC00168) gave ethical approval for this work(HREC/18/QPCH/179), with subsequent authorisation to conduct the project at the Sleep Assessment Unit at the Sunshine Coast University Hospital (SCUH) (QSC/40666).

